# Identifying subtypes of youth suicidality based on psychopathology: alterations in genetic, neuroanatomical and environmental features

**DOI:** 10.1101/2024.04.13.24305772

**Authors:** Xinran Wu, Lena Palaniyappan, Laura van Velzen, Gechang Yu, Huanxin Fan, Liu Yu, Benjamin Becker, Wei Cheng, Xingming Zhao, Jianfeng Feng, Barbara J. Sahakian, Trevor W. Robbins, Gunter Schumann, Lianne Schmaal, Jie Zhang

**Affiliations:** Institute of Science and Technology for Brain-Inspired Intelligence, Fudan University, Shanghai 200433, PR China; Key Laboratory of Computational Neuroscience and Brain Inspired Intelligence, Fudan University, Ministry of Education, PR China; Douglas Mental Health University Institute, Department of Psychiatry, McGill University, Montreal, QC, Canada; Department of Psychiatry, Schulich School of Medicine and Dentistry, Western University, London, ON, Canada; Robarts Research Institute, University of Western Ontario, London, ON, Canada; Department of Medical Biophysica, Schulich School of Medicine and Dentistry, Western University, London, ON, Canada; Orygen, The National Centre of Excellence in Youth Mental Health, Parkville, VIC, Australia; Centre for Youth Mental Health, The University of Melbourne, Parkville, VIC, Australia; Department of Medicine and Therapeutics, The Chinese University of Hong Kong, Prince of Wales Hospital, 999077, Hong Kong SAR of China; State Key Laboratory of Brain and Cognitive Sciences, The University of Hong Kong, Hong Kong SAR; Shanghai Center for Mathematical Sciences, Shanghai, 200433, People’s Republic of China; Department of Computer Science, University of Warwick, Coventry, CV4 7AL, UK; Collaborative Innovation Center for Brain Science, Fudan University, Shanghai, 200433, People’s Republic of China; Fudan ISTBI—ZJNU Algorithm Centre for Brain-inspired Intelligence, Zhejiang Normal University, Jinhua, People’s Republic of China; Department of Psychiatry, Cambridge Biomedical Campus, University of Cambridge, Cambridge, UK; Department of Psychiatry and Behavioural and Clinical Neuroscience Institute, University of Cambridge, Cambridge, United Kingdom; The Centre for Population Neuroscience and Stratified Medicine (PONS), ISTBI, Fudan University, Shanghai, China; PONS Centre and SGDP Centre, Institute of Psychiatry, Psychology and Neuroscience, King’s College London, London, UK; PONS Centre, Charite Mental Health, Dept. of Psychiatry and Psychotherapie, CCM, Charite Universitaetsmedizin Berlin, Berlin, Germany

## Abstract

One of the most complex human behaviours that defies singular explanatory models is suicidal behaviour, especially in the youth. A promising approach to make progress with this conundrum is to parse distinct subtypes of this behaviour. Utilizing 1,624 children with suicidal thoughts and behaviors (STBs) and 3,224 healthy controls from the ABCD Study, we clustered children with STB based on thirty-four cognitive and psychopathological measures which capture suicide-related risk-moderating traits. Environmental and genetic risk factors, as well as neuroanatomical characteristics of each subtype, were then compared with controls. We identified five distinct STB subtypes, each revealing unique neuroanatomy, environmental/genetic risks, and persistence patterns. Subtype 1 (Depressive, 9.6%) exhibited the most severe depressive symptoms. Subtype 2 (Externalizing, 20.1%) displayed anatomical and functional alterations in frontoparietal network and increased genetic risk for ADHD. Subtype 3 (Cognitive Deficit, 20.4%) demonstrated lower cognitive performance and widespread white-matter deficits. Subtype 4 (Mild Psychotic, 22.2%) presented higher prodromal psychotic symptoms, often unnoticed by parents. Subtype 5 (High Functioning, 27.6%) showed larger total brain volume, better cognition, and higher socio-economic status, contrasting subtypes 1-4. Only Subtypes 1 and 2 demonstrate persistent STB features at the 2-year follow-up. Our results suggested that youth suicidal behaviour may result from several distinct bio-behavioral pathways that are identifiable through co-occurring psychopathology, and provide insights into the underlying neural mechanisms and corresponding intervention strategies.

## INTRODUCTION

Despite advances in the assessment and treatment of suicidal thoughts and behaviors (STBs), suicide is still one of the leading causes of mortality worldwide, especially for youths ^1^. The incidence of suicidal attempt peaks in mid-adolescence, and is the second major cause of death among young people between the ages of 10 and 14 ^2^. Recent COVID-19 pandemic further heightened the risk of suicide, especially among children and adolescents, with a 10% increase in emergency room visits for STBs ^3^.

There is a call for fine-grained phenotyping of suicidal behaviour, as a unified approach searching for final common pathway towards youth suicide has not been fruitful to date ^4^. If there are multiple pathways, each with its own initial mechanistic point (or diathesis) of deviation and distinct trajectories shaped by the meso-system (school, family, peers), then blanket interventions we offer to STBs that do not address all of these causal pathways will only have small effects; such small effects are precisely what has been reported in a recent meta-analysis ^5^.

There are different lines of evidence supporting the notion that STB roots in multiple potential subtypes. First, even well-known risk factors (such as trauma, malignant stress) do not result in STBs in all those who are exposed ^6^; there appears to be a distinct set of individuals who may be more prone (a model of multiple diatheses, multiple stressors) ^7^. Second, this variation does not appear to be random; qualitative differences based on sociological, e.g., Durkheim’s description of typology ^8^, or psychological domains are well noted among adults with suicide risk ^9–11^. Third, there is well founded empirical evidence for grouping STBs with distinct clinical features as subtypes. For example, based on observations pertaining to stress-reactivity, Bernanke et al. adopted temporal stability to classify STBs into stress-responsive (intermittent) and non-stress-responsive (persistent) subtypes ^4^. As a result, researchers have begun to consider STB as a final expression stemming from different subtypes with multiple separate pathological changes ^4^. Reliably separating these subtypes will help to better understand the neural mechanisms behind suicide and develop better tailored prevention and intervention efforts.

According to the stress-diathesis model, suicidal behavior can be viewed as a result of interactions between two main factors: acute stressors or proximal risk factors, and diathesis or distal factors ^12^. Diathesis refers to a set of relatively stable traits associated with suicide, and heterogeneity of diathesis constitutes the distinct coping styles in the face of acute stressors and therefore can serve as a valuable feature for identifying distinct suicide subtypes. On this basis, previous researchers have investigated the heterogeneity of STB-related diatheses, and some partially reproducible STB subtypes, such as the impulsive type, depressive/anxious type ^13,14^, have been obtained. However, these are not enough to determine usable child STBs subtypes. First, most previous studies were based on adults, while child STB shows distinct features, such as being more impulsive ^15^ and greater susceptibility to social bullying or peer pressure ^16^. Second, due to its transdiagnostic nature, characterizing the complete heterogeneity of STB requires multidimensional psychopathological and cognitive measurements usually ignored. Third, only clinical symptoms were investigated in previous studies, but the underlying neural mechanisms and genetic/environmental risk factors were not fully examined. These limitations could be overcome by new, large-sample multidimensional clustering studies to provide more clinically significant subtypes for child STBs and by exploring the neural mechanisms of different subtypes crucial for targeted intervention.

In this study, a large child sample (1,624 children with suicidality, 3,228 controls, 9-10 years old) from the Adolescent Brain Cognitive Development (ABCD) study was used to investigate heterogeneity of children suicidal thoughts and behaviors. In this study, we selected 34 psychological variables based on the classic stress-diathesis model ^7^ in the field of suicide research, which were input into a data-driven clustering approach to identify subtypes of child suicidality. We furthermore compared demographic, environmental, neuroimaging and genetic characteristics, and development of suicide symptoms over time between subtypes and healthy controls, which help substantiate the distinct subtypes identified and provide comprehensive fingerprints that may offer clinically valuable information.

## RESULTS

### Study population

11,878 children recruited from 22 sites from The Adolescent Brain Cognitive Development (ABCD) study (version 3.0). Suicidal Thoughts and Behaviors (STBs) and Healthy Control (HC) groups were identified using the Kiddie Schedule for Affective Disorders and Schizophrenia Present and Lifetime Version (K-SADS) at baseline (10-years old), consisting of both parent-report (K-SADS-P) and youth self-report (K-SADS-Y). The STB group includes individuals with suicidal ideation, attempts, or behavior reported in either K-SADS-P or K-SADS-Y (N_STB_ = 1,624). The HC group comprises of those without any reported diagnoses in either K-SADS-P or K-SADS-Y at baseline (N_HC_ = 3,228).

### Five distinct subtypes of suicidality identified in children

We selected the features required for the cluster analysis based on the widely accepted stress-diathesis model of suicide ^7^. According to the stress-diathesis model, distress factors related to suicide encompass the following domains:1) subjective distress and attentional bias to negative stimuli; 2) decision-making defects leading to impulsive and emotional behavior; 3) cognitive impairment, such as learning and memory problems; 4) social distortions. We utilized 34 normalized cognitive and psychopathological variables from 5 questionares in ABCD which captured these suicide-related diathesis (see Methods for details) to cluster children with STBs. K-means clustering with bootstrap adopting Euclidean distance was employed (number of clusters K=2-10), and an optimal K was selected by gap statistic ^17^. 5 clusters/subtypes were finally identified (gap statistic=0.81, see SM Fig. 2):

1. Subtype 1 (depressive type, n=156, 9.6%) had the highest Depression and Anxiety score, and Total Problem score of CBCL, and the highest morbidity rate of general anxiety disorder (GAD, 54.5%), phobia (62.8%) and ADHD (55.8%), see Table 1.
2. Subtype 2 (externalizing type, n=327, 20.1%) had high externalizing symptom score in CBCL, especially for Attention Problems, ADHD scale and oppositional defiant disorder (ODD) scale, and the highest morbidity rate of ODD (57.5%) and CD (conduct disorder, 23.5%).
3. Subtype 3 (cognitive deficit type, n=331, 20.4%) had the most cognitive impairment, and the lowest behavioral inhibition/activation (BIS/BAS) score.
4. Subtype 4 (mild psychotic type, n=361, 22.2%) had the highest psychotic-like symptoms (PPS) and impulsivity (UPPS and BIS/BAS).
5. Subtype 5 (high functioning type, n=449, 27.6%) demonstrated no difference or even slightly better cognitive ability than HC.

**Figure 1.**
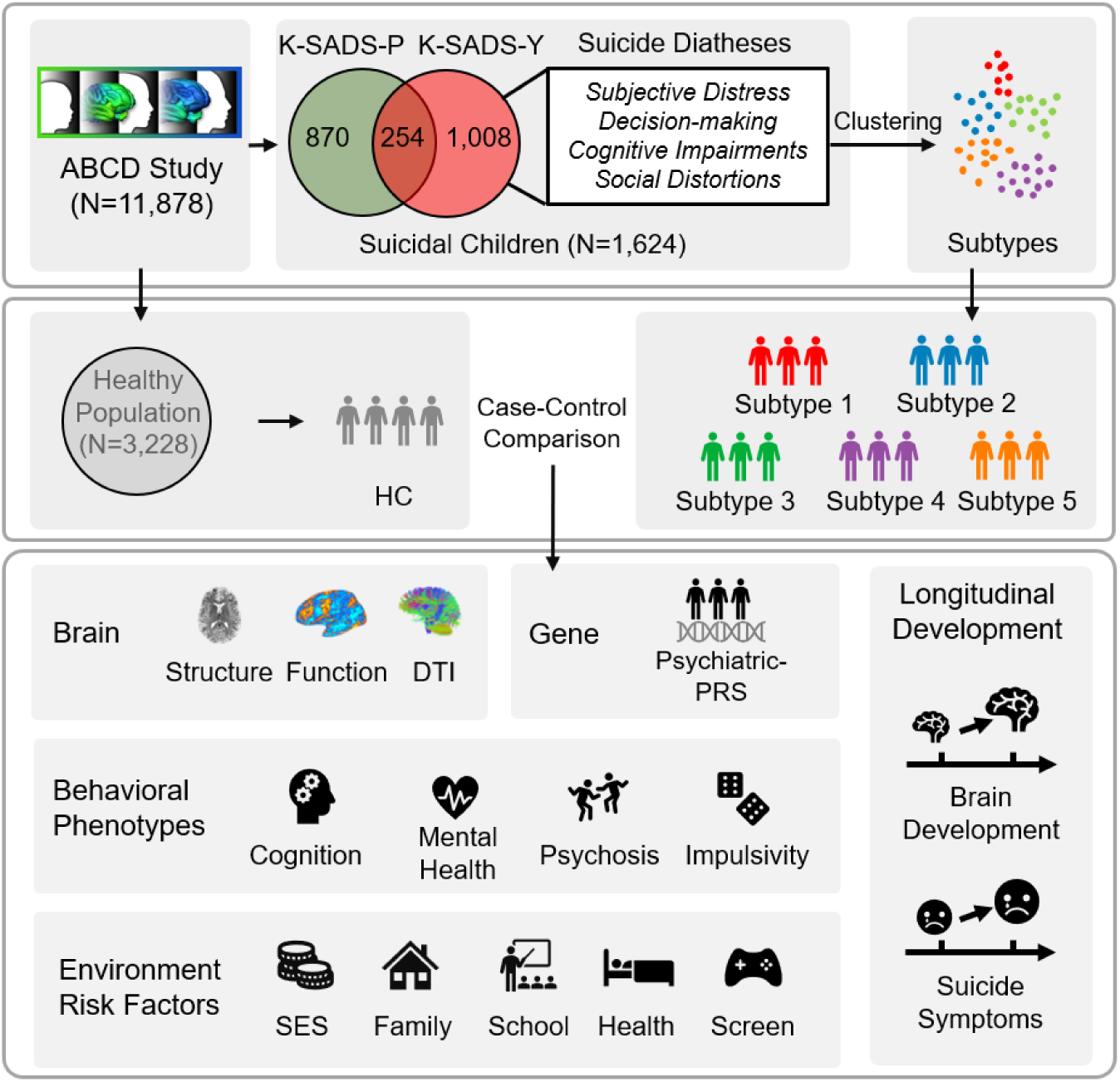
Workflow of the whole study. 11,878 children from ABCD study were used to screen out 1,624 children with suicidal ideation, attempt or behavior on the parent- or youth-report K-SADS suicidality items. We used multiple behavioral phenotypes (diatheses) about subjective distress, decision-making deficits, cognitive impairments, and social distortions. as features. Clustering algorithm was used to obtain 5 subtypes of suicidal children. Finally, we compared differences in brain structure/connectivity measures, environmental factors, genetic risk measures, and development between these five subtypes and between them and a healthy control group of 3,228 children. K-SADS-P, Parent-reported Kiddie Schedule for Affective Disorders and Schizophrenia. K-SADS-Y, Youth-reported Kiddie Schedule for Affective Disorders and Schizophrenia. Note: 1,624 suicidal children were composed of 870 children with suicidality defined by K-SADS-P and 1008 children with suicidality defined by K-SADS-Y, among which 254 overlapped; HC, healthy control group.

**Figure 2.**
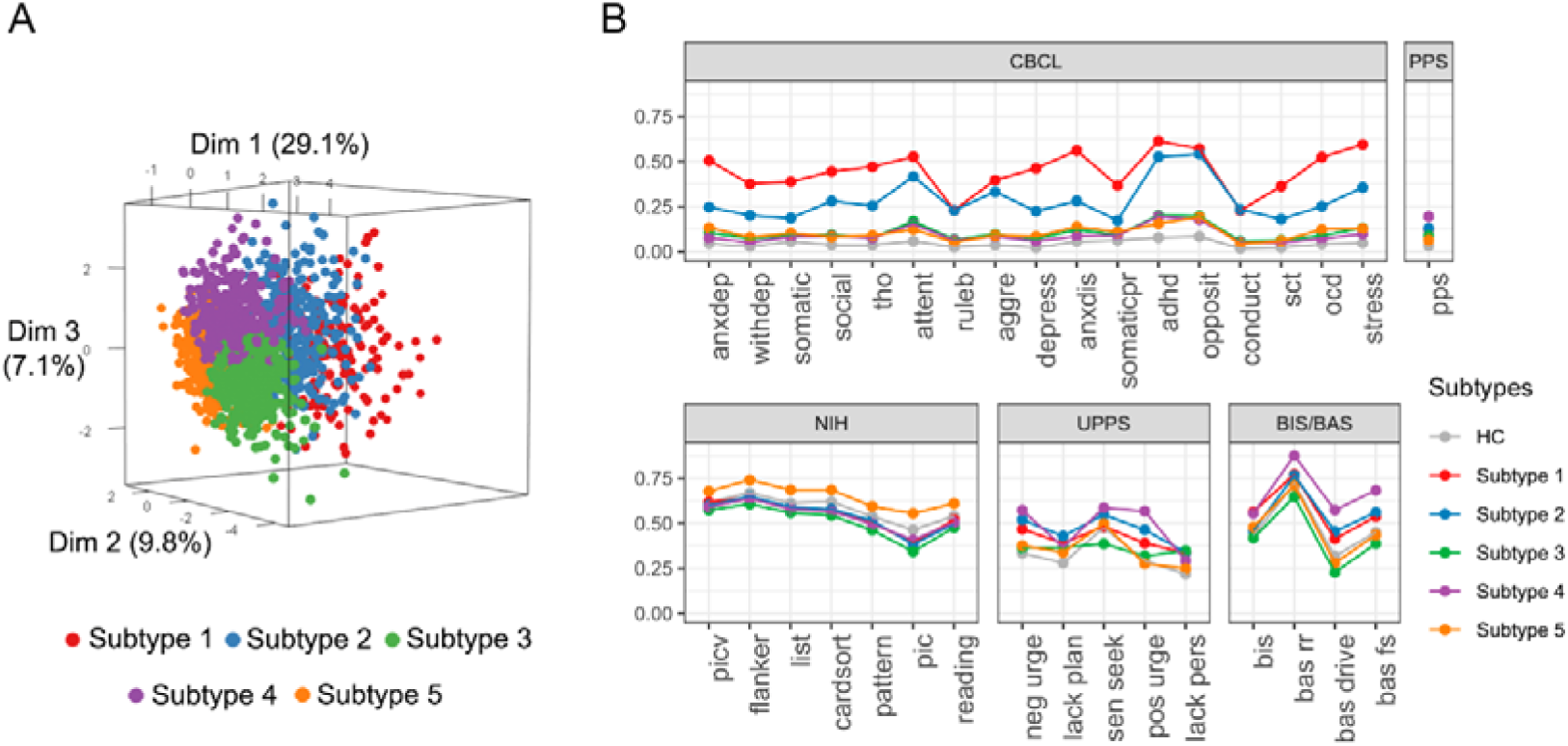
Full-spectrum of Behavioral phenotypes of STB subtypes. (A) three-dimension scatter plot of suicidality subtypes. Each dot represents a subject, and the three dimensions (Dim 1, Dim 2, Dim 3) represent the first 3 principal components of PCA extracted by 34 features for clustering. The numbers in brackets represent the amount (%) that the PC explains the total variation. The color of the dots represents the subtype. (B) profiles for each subtype and healthy control group (HC). All features were normalized between 0 and 1. The full names of the abbreviations in figure can be found in SM Table 1.

**Table 1.** Demographic and clinical characteristics of healthy control participants & the five subtypes at baseline. The numbers in parentheses indicate the proportion of children with this attribute in each subtype. AIAN, American Indian/Alaska Native; NHPI, Native Hawaiian and other Pacific Islander.

|  | HC | Subtype 1 | Subtype 2 | Subtype 3 | Subtype 4 | Subtype 5 | p |
| --- | --- | --- | --- | --- | --- | --- | --- |
| N | 3228 | 156 | 327 | 331 | 361 | 449 |  |
| Age by Months (mean) | 119 | 119.86 | 119.35 | 116.27 | 117.44 | 121.18 | <0.001 |
| Sex=Male (%) | 1476(45.7) | 93(59.6) | 228(69.7) | 185(55.9) | 220(60.9) | 251(55.9) | <0.001 |
| Race (%) |  |  |  |  |  |  | <0.001 |
| White | 2089(64.7) | 93(59.6) | 192(58.7) | 183(55.3) | 204(56.5) | 335(74.6) |  |
| Black | 475(14.7) | 24(15.4) | 61(18.7) | 55(16.6) | 76(21.1) | 14(3.1) |  |
| Asian | 99(3.1) | 1(0.6) | 1(0.3) | 7(2.1) | 7(1.9) | 26(5.8) |  |
| Mixed | 349(10.8) | 29(18.6) | 60(18.3) | 61(18.4) | 40(11.1) | 61(13.6) |  |
| Other | 145(4.5) | 6(3.8) | 11(3.4) | 17(5.1) | 22(6.1) | 10(2.2) |  |
| AIAN/NHPI | 26(0.8) | 2(1.3) | 0(0) | 2(0.6) | 2(0.6) | 2(0.4) |  |
| Pubertal (mean) | 1.89 | 2.05 | 1.96 | 1.89 | 1.97 | 1.9 | 0.065 |
| Married or living together (%) |  |  |  |  |  |  | <0.001 |
| yes | 2507(77.7) | 91(58.3) | 191(58.4) | 225(68) | 249(69) | 374(83.3) |  |
| no | 699(21.7) | 65(41.7) | 132(40.4) | 102(30.8) | 110(30.5) | 74(16.5) |  |
| BMI (mean) | 18.94 | 19.49 | 19.04 | 18.78 | 19.13 | 18.32 | 0.007 |
| Income (mean) | 2.19 | 1.8 | 1.83 | 2.03 | 2 | 2.4 | <0.001 |
| Education (mean) | 3.76 | 3.65 | 3.47 | 3.61 | 3.59 | 4.17 | <0.001 |

**Table 2.**
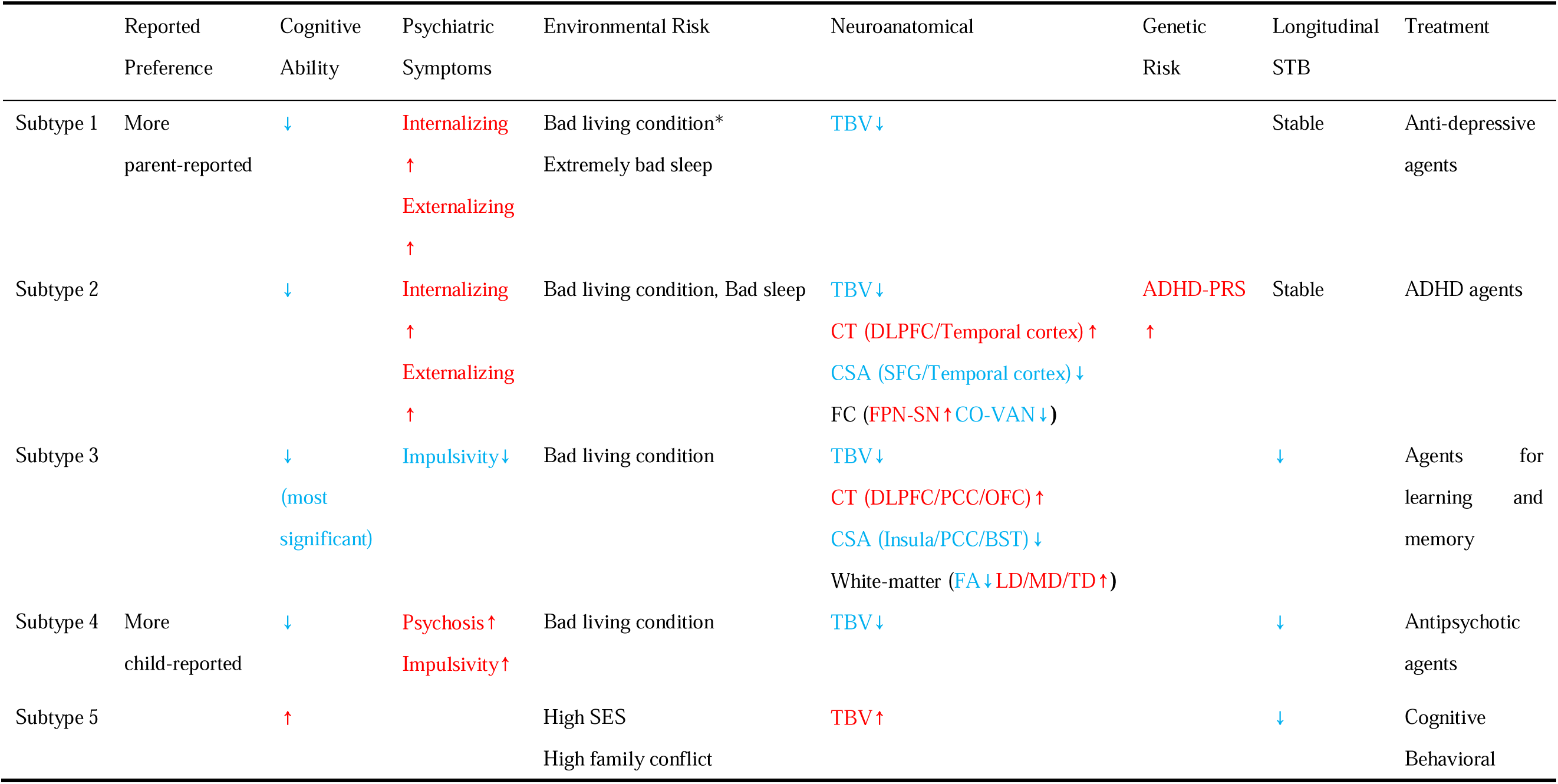
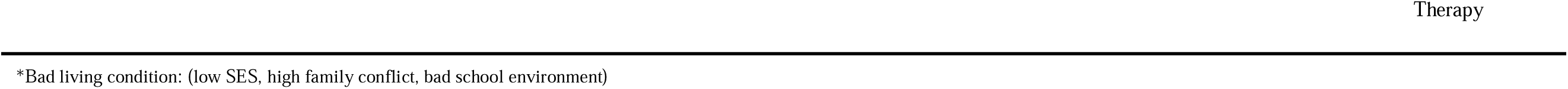
Summary of the five STB subtypes. The red up arrow indicates significant increase relative to healthy controls, and the blue down arrow indicate significant decrease. TBV, total brain volume; CT, cortical thickness; CSA, cortical surface area; FC, functional connectivity; DLPFC, dorsolateral prefrontal cortex; OFC, orbitofrontal cortex; BST, banks of superior temporal sulcus; PCC, posterior cingulate cortex. FPN, frontoparietal network. SN, salience network. CO, cingulo-opercular network. VAN, ventral attention network.

One-way ANOVA (Analysis of Variance) was used to analyze the differences in continuous variables between the various subtypes and the healthy control group, while chi-square tests were used to analyze the differences in categorical variables. In general, all subtypes had higher CBCL and PPS scores than HC. Subtypes 1 and 2 had higher proportion of prepared suicidal behavior than the other 3 subtypes (Fig. 2B and SM Table 3-6). To determine if there are systematic differences between parent and children reports, we compared the frequency of parent- and child-reported STBs for all subtypes. Subtype 1 (depressive) were more often reported by parents (K-SADS-P: 19%, K-SADS-Y: 10%), while subtype 4 (mild psychotic/impulsive) were more reported by children themselves (K-SADS-P: 12%, K-SADS-Y: 23%), suggesting it was either difficult for parents to detect STBs when children have psychotic/impulsive features or for the children with these issues to articulate their suicidal thoughts to parents (Fig. 4A and Table 1). The proportion of male was higher in all subtypes than that in HC, with subtype 2, the externalizing type (69.7%, p<0.001) being the most prominent (Fig. 4B and Table 1). The proportion of white and black children with STBs were lower and higher than that of HC, respectively, for all subtypes except subtype 5, see Fig. 4C.

### Social and environmental risk profiles of different subtypes

Risk factors, such as adverse environmental factors and physical health, play a pivotal role in precipitating suicidality among children ^18,19^. We delved into inter-group differences using one-way ANOVA to identify potential risk factors for distinct subtypes, encompassing Socioeconomic Status (income, parental education, neighborhood safety), Family Environment (parental marital status, family conflict, etc.), School Environment, Health (sleep quality, sports involvement), and time spent using electronic screen devices (SM Table 1-2).

5 STB subtypes demonstrate common and unique risk factors. All subtypes had significantly more severe sleep disturbances, more family conflict, lower acceptance score from parents, less school involvement, and more school disengagement than HC (Table 1 and SM Tables 5-6). There were significant inter-group differences in parental education (F_4842_=18.1, p=7.74e-18) and household income (F_4427_=26.9, p=6.56e-27), with subtype 1-4 showing lower values than HC (p<0.001), while subtype 5 showed higher values than HC (parental education: diff_5-0_=0.41, p=2.1e-8; household income: diff_5-0_=0.2, p=3.7e-5). The parents divorce rate in all 5 subtypes was higher than that in HC. To summarise, subtypes 1-4 faced a harsher home environment than HC, while the high-functioning subtype had better living conditions despite family conflicts and school-level problems.

### Neurodevelopmental alterations in different STB subtypes

We employed linear mixed models (LMMs) to evaluate the changes in gray matter structure, white matter microstructure, and functional connectivity features of each subtype relative to the healthy control (HC) group, which allowed us to identify both common and distinct multimodal neuroimaging changes across the subtypes. In terms of commonalities, all subtypes, except the high-functioning subtype, exhibited a decrease in Total Brain Volume (TBV): depressive type (t_4285.95_=-2.54, p=0.011), externalizing type (t_4259.19_=-5.65, p=1.71e-8), cognitive deficit type (t_4271.41_=-3.52, p=4.39e-4) and mild psychotic type (t_4249.23_=-3.52, p=4.39e-4). High functioning type showed slightly higher TBV compared to controls (t_4256.14_=2.18, p=0.0296). Thus, the TBV pattern is similar to the socio-environmental risk profile.

We also identified subtype-specific structural and functional alterations (Fig. 3B-D). For brain structure, the externalizing type showed larger cortical thickness (CT) in dorsolateral prefrontal cortex (DLPFC), middle and superior temporal, isthmus, posterior and middle cingulate, and significantly lower cortical surface area (CSA) in superior / inferior temporal and temporal pole, left precentral and superior parietal cortex than HC (FDR q<0.05, TBV regressed). Cognitive deficit type shared some but not all of these features; this group showed significantly higher CT in DLPFC, medial orbitofrontal, precuneus, bilateral isthmus cingulate, postcentral/paracentral and cuneus, and lower CSA in banks of superior temporal sulcus (BST), posterior cingulate cortex and insula than HC (FDR q<0.05). High functioning type showed lower CT of the left precuneus (t_4404.02_=-3.49, p=4.91e-4) and smaller CSA of right bank of superior temporal sulcus (t_4365.81_=-3.59, p=3.29e-4), see Fig. 2B. This indicates mre widespread evidence of delayed maturation of transmodal (especially frontotemporal) areas as we move from subtype 5 (high functioning) to 3 (cognitive deficit) and 2 (externalizing type).

**Figure 3.**
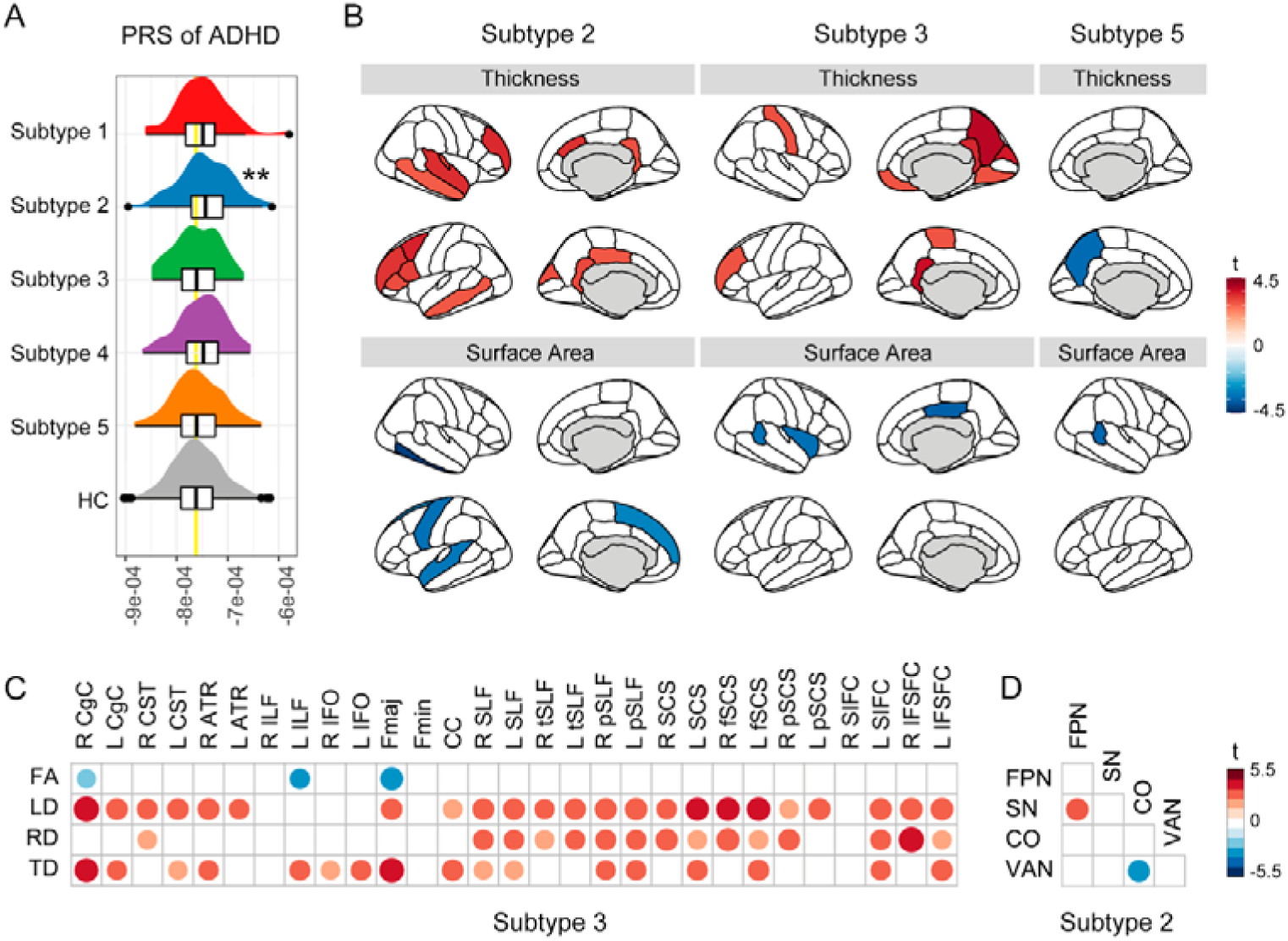
Difference in brain and PRS between STB subtypes and controls. (A) Differences in the polygenic risk score (PRS) of ADHD between suicidal subtypes and controls. The bright yellow line indicates the mean ADHD-PRS of the HC group. Only PRS for Subtype 2 was significantly greater than HC (two asterisks denoting p<0.01). (B) Differences in cortical surface area and cortical thickness between suicidal subtypes and controls. (C) Differences in white matter fiber microstructure between subtype 3 and controls. FA, fractional anisotropy. MD, mean diffusivity. LD, longitudinal diffusivity. TD, transverse diffusivity. Only LMM-based t values for all significant tracts are shown in B-D (FDR q<0.05). (D) Differences in resting-state functional connectivity between subtype 2 and controls. FPN, frontoparietal network. SN, salience network. CO, cingulo-opercular network. VAN, ventral attention network.

For DTI, only the cognitive deficit type showed significant but widespread alterations (lower FA and higher LD/RD/TD than HC). The most significant changes (FDR q<0.01) were in right cingulate cingulum (CgC, LD: t_3893.68_=4.22, p=2.46e-5; TD: t_3836.67_=3.73, p=1.93e-4), right inferior to superior frontal tract (IFSFC, LD: t_3893.68_=4.18, p=2.96e-5), forceps major (Fmaj, TD: t_3888.66_=3.95, p=7.94e-5) and bilateral frontal superior corticostriate (fSCS) (LD of left fSCS: t_3865.54_=3.65, p=2.62e-4; LD of right fSCS: t_3876.67_=3.63, p=2.92e-4). In addition, corticospinal, thalamic radiations, longitudinal fasciculus, frontal occipital fasciculus and corpus callosum also showed significant differences (all FDR q<0.05, Fig. 2C).

For functional connectivity, only the externalizing type showed lower FC between frontoparietal network (FPN) and salience network (SN) (t_3427.72_=-3.36, p=7.86e-4), and higher FC between cingulo-opercular network (CO) and ventral attention network (VAN) (t_3549.1_=3.28, p=1.06e-3) than HC (Fig. 2D).

In addition to detecting differences between each subtype and HC group, our model also examined the developmental effect (the changes of brain features for all group from baseline to 2-year follow-up) and interaction effects (the age-related brain changes of each subtypes). No significant interactions were found, suggesting that the brain development of each subtype (compared to HC) did not differ significantly from ages 10 to 12 (SM Table 7-9), and the distinct patterns in structure and connectivity reported above are mostly established prior to the baseline scan (before age 9). Since the average developmental effects are unrelated to the main theme, they are not elaborated here. (see Supplemental Materials).

### Genetic risk for psychiatry in different STB subtypes

To examine the potential psychopathological genetic risk of 5 suicidality subtypes, polygenic risk scores (PRSs) of six psychiatric illnesses (ADHD, BP, MDD, OCD, PTSD and schizophrenia) were calculated and compared between 5 subtypes and HC using one-way ANOVA. PRSs were calculated by PRS-CS ^20^using GWAS summary statistics of the above six mental disorders from the Psychiatric Genomics Consortium (PGC) on 4,933 unrelated European-ancestry individuals. Only the externalizing type showed higher ADHD-related PRS than HC (F_1953_= 4.84, p=0.0002; post-hoc: p=4.8e-4, one-way ANOVA) see Fig. 2A and SM Table 10-11. PRSs of MDD, OCD, PTSD, schizophrenia and BIP did not show significant difference between each subtypes and HC.

### Longitudinal persistence of STBs

To determine which subtype have chronic suicidality, we defined the STB score as the total number of reporting “yes” in K-SADS items about suicidal thoughts, attempts, behaviors and self-injury (separately for K-SADS-P and K-SADS-Y), and conducted a repeated measures 5*2 ANOVA, which examined the main effects of subtypes and age (baseline and 2-year follow-up), and interaction effect (Supplemental Methods). All 5 subtypes showed declining STB scores with age. A significant subtype-age interaction was observed in K-SADS-Y STB scores (F_930_=3.47, p=0.008). Post-hoc analysis indicated no significant age-related decrease in STB scores for depressive and externalizing subtypes (t=-1.94, p=0.06 and t=-2.5, p=0.013), while the other subtypes showed significant age-related reductions (SM Table 12-13).

### Validation analysis

To assess the reproducibility of the clustering results, validation analysis was performed. First, split-half analysis (split the data into two halves and repeated the clustering analysis) replicated the main results (SM Fig. 2 and SM Table 14). Second, clustering was performed separately using only parental or child reported STBs. 6 subtypes were identified in youth-reported STBs (N=1,007), including all 5 subtypes previous identified. Five subtypes were found in parent-reported STBs, replicating all subtypes except the mild psychosis type (SM Fig 3), highlighting that this subtype was most likely to be missed by parents.

## DISCUSSION

This study used a large transdiagnostic sample of 1,624 children to identify meaningful subtypes among preadolescents with suicidal thoughts and behaviors (STBs) based on multidimentional suicide-related risk-moderating diatheses. We noted that the largest proportion of suicidal children have higher socioeconomic status, brain volumes and cognitive abilities than healthy peers, and show relatively transient STB. The next commonly occurring subtype is prone to psychotic features, and their suicidal thoughts are often unknown to their parents. The other 3 subtypes exhibit psychopathology in depressive, externalizing or impaired cognition domains, and show lower brain volumes, regional cortical surface area while higher cortical thickness than healthy peers, indicating a maturational lag that likely preceded in onset before the age 9. Of these, the externalizing subtype also showed abnormal resting-state functional connectivity among cognitive control networks and higher genetic risk for ADHD, while the cognitively impaired subtype demonstrated widespread white-matter microstructure alterations, indicating individual-level neural contributors to the phenotypes. Interestingly, parental conflicts and school disengagement was high in all 5 subtypes, indicating a shared pattern of socio-familial dysfunction in these domains.

When considering the meaning of the observed subtypes, it is essential to revisit the notion of heterogeneity. In general, a large range of deviation around a prototypical profile contributes to heterogeneity in biosystems. But in some cases (like STBs), a single prototype does not exist; multiple prototypes (subtypes) may explain the lack of unified mechanisms and treatment response trajectories ^21^. This heterogeneity may explain why a singular brain-centric model of youth STBs has been elusive to date ^7^. For example, when reduced thickness of left superior temporal sulcus was observed without considering subtypes in a prior ABCD-based analysis, the effect size was small ^22^; another machine learning study using the same ABCD data also reported low contribution of neuroimaging features to suicidal behavior prediction ^23^, suggesting treating all suicidal children as one homogeneous group prevents identification of sensitive STBs biomarkers.

Our observations not only show consistency with previous studies, but also add neuro-anatomical (morphometric and connectivity-based) and genetic validation to early reports of STB, taking us closer to explaining the mechanisms and constructs that contribute to these subtype patterns. There are some recurring subtypes reported both in our study and previous adult STB studies, such as non-stress-responsive (persistent suicidal thoughts and depression) and stress-responsive types (high impulsivity) ^4,24^, matching our depressive subtype and externalizing subtype, respectively. Moreover, the subtypes we identified substantiate gender-related distinctions in STB patterns observed in previous research ^25^. The subtype 2 we identified, exhibiting externalizing issues and persistent suicidality, is predominantly composed of males (69%, Table 1), aligns with prior findings in male students^26^. In contrast to our results where a greater proportion of males were observed in all subtypes (Table 1), earlier studies reported higher STB rate among female adolescents than males ^27^, potentially due to the 10-year-old children in our cohort had not yet entered adolescence, thus masking the hormonal influences on female suicidality.

Our neuro-anatomical and genetic discoveries pertaining to these subtypes rest firmly on prior findings, but provide novel directions to understand youth suicidality. Firstly, all subtypes except high functioning type showed lower TBV than HC. Lower TBV may be associated with premature excessive synaptic pruning triggered by environmental stress in childhood ^28^ and impaired intelligence ^29,30^, consistent with our findings (Fig 4C). High functioning type, by contrast, exhibits larger TBV than HC, suggesting TBV is more likely to be determined by better living conditions, and contributes to cognitive compensation in the presence of factors co-occur with youth suicidality (parental conflicts, poor sleep and school disengagement). Secondly, externalizing and cognitively impaired suicidal youth (subtype 2 and 3) share neuroanatomical abnormalities in emotional regulation circuit (e.g., increased thickness of DLPFC and PCC), one of the core substrates of the suicidal brain ^31^. DLPFC supports top-down emotional inhibition ^32^, and PCC regulates emotions via attention deployment ^33^. Therefore this results underscore the importance of subtype-specific targeting of neural systems, rathr than focusing on general brain health, when attempting to design empirically informed interventions for youth STBs.

**Figure 4.**
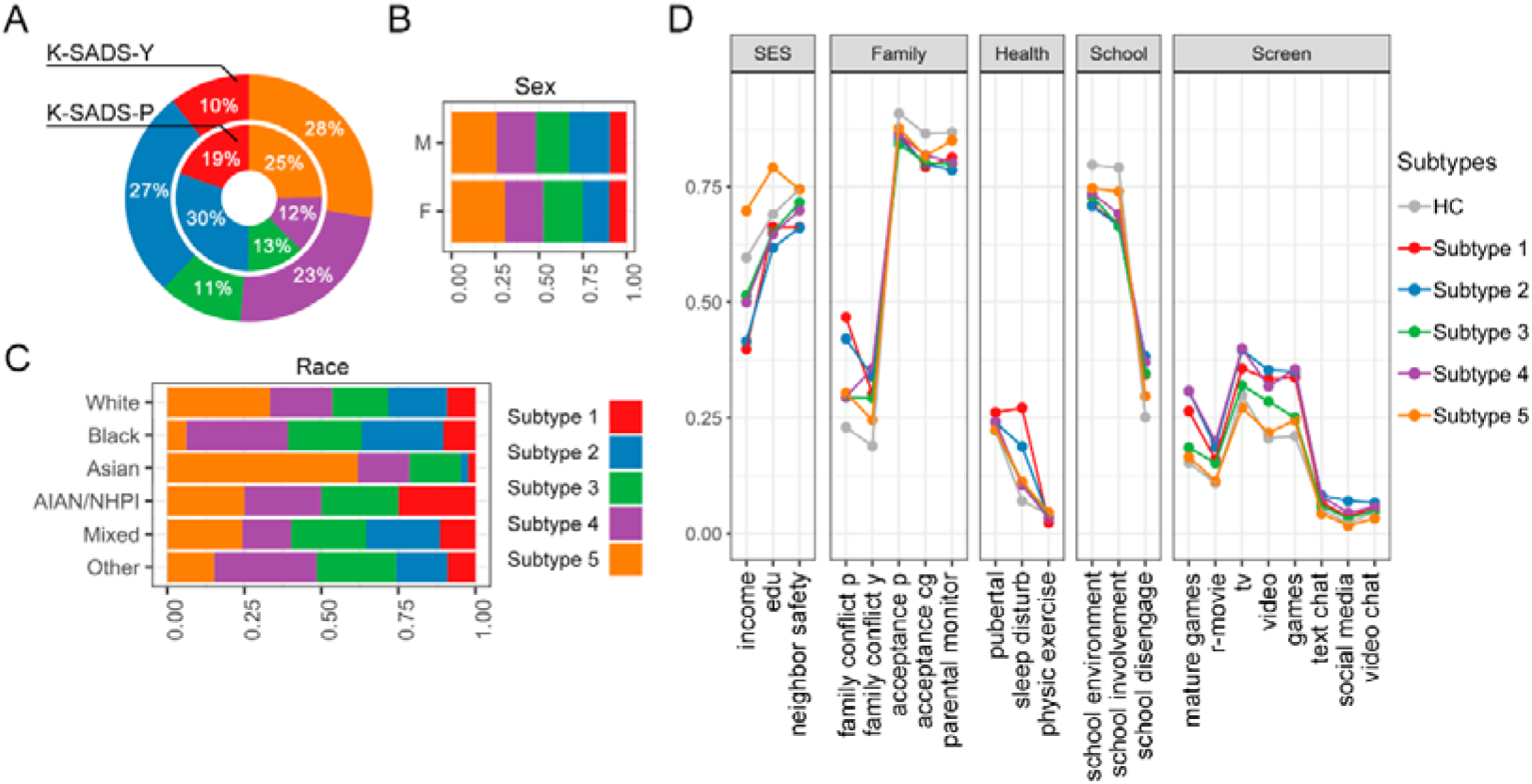
Demographic and environmental variables. (A) The proportion of suicidal children of different subtypes in parent-reported and youth-reported suicidality. The outer circle represents the youth-reported and the inner circle represents the parent-reported suicidality from Kiddie Schedule for Affective Disorders and Schizophrenia (K-SADS). K-SADS-P, parent-report K-SADS. K-SADS-Y, youth-report K-SADS. (B-C) The proportion of subtypes of STB children by sex/race. The x-axis length of each bar is 1, representing the people with a certain attribute in all suicidal individuals. Different colors represent the relative proportions of 5 subtypes. M, Male; F, Female. AIAN, American Indian/Alaska Native; NHPI, Native Hawaiian and other Pacific Islander. (D) Profiles of environmental variables for each subtype and healthy control group (HC). SES, Socioeconomic status. FES, Family Conflict Subscale of Family Environment Scale. CRPBI, Child Report of Behavior Inventory. SDS, Sleep Disturbance Scale. SRPF, School Risk and Protective Factors. The full names of the abbreviations can be found in SM Table 1.

Different suicide subtypes also have unique neuroanatomical bases. The externalizing subtype displays abnormalities in control and attention-related networks: salience and cingulo-opercular networks are both centered on insulathat was involved in interoception and prediction error signaling ^34^, while disconnection between FPN and VAN likely reflects the failure to integrate top-down ^35,36^ and bottom-up sources of attentional regulation ^37,38^. This suggested that externalizing type has dysregulation in cognitive control. Moreover, high genetic risk of ADHD in this subtype is a well-known risk factor for suicide, likely related to impaired attention and diminished ability to regulate emotions by attentional control ^39,40^. In comparison, the cognitive deficit subtype shows wide microstructural white matter defects, e.g., in superior longitudinal fibers associated with intellegence^41^, which may affect problem solving and increase their stress and suicide risk ^42^.

Identifying consistent subtypes among suicidal youth allows for systematic exploration of treatment matching and tailored care. At present, the interventions offered for suicidality in youth are limited in scope and evidence base and are not mapped onto the developmental context^43^. While system-level interventions like school-based programs show potential ^44^, their long-term effects are uncertain and are inconsistently implemented due to lack of replication and weak effect size. Our clustering utilizes assessable symptoms and behaviors, applicable across various settings (schools, primary care units, social agencies), without the need for diagnosis, scans, or genetic data. This approach provides a foundation for estimating treatment benefits in stratified STB subtypes in ongoing prevention/intervention studies. For example, the externalizing type shows abnormalities in control and attention networks and a genetic risk of ADHD, therefore might benefit from psychostimulant medication for ADHD treatment (such as methylphenidate and amphetamine derivatives), which enhances the inhibitory control and have proven to be associated with reduced risk of suicide attempts in ADHD patients ^45,46^ or children with externalizing symptoms ^47^.

### Strengths & Limitations

Our study has several strengths: In a data-driven analysis to resolve subtypes of STBs in children of 9-10 years old using a large transdiagnostic cohort, we conducted multilevel validation using genetic, MRI, demographic, symptom and cognitive data. Secondly, we examined long term persistence of STBs using 2-year follow-up data. This study also has limitations. First, the age range in ABCD study v3.0 is from 9 to 12, so individuals with STB in late adolescence is not studied. Other limitations include the absence of a severity measure of suicidality and the therapeutic outcomes of each subtype, Future investigations are needed to determine the most suitable treatment for each subtype.

## METHODS

### Definition of suicidal thoughts and behaviors (STBs) in children

Suicidal Thoughts and Behaviors (STBs) and Healthy Control (HC) groups were identified using the Kiddie Schedule for Affective Disorders and Schizophrenia Present and Lifetime Version (K-SADS) at baseline (10-years old), consisting of both parent-report (K-SADS-P) and youth self-report (K-SADS-Y). The STB group includes individuals with suicidal ideation, attempts, or behavior reported in either K-SADS-P or K-SADS-Y (N_STB_ = 1,624). We combined parent-reported (N=870) and child-reported STBs (N=1,008). Because there is small overlap between the parent and child reports (N=259). Moreover, some children may hesitate to disclose STBs to interviewers / caregivers, thus relying solely on parent reports is incomplete. The HC group comprises of those without any reported diagnoses in either K-SADS-P or K-SADS-Y at baseline (N_HC_ = 3,228).

### Detecting subtypes in children STBs by multi-dimensional suicidal diatheses

According to the stress-diathesis model ^7^, the suicide-related risk-moderating traits involves 4 aspects: subjective distress, decision-making deficits, cognitive impairments, and social distortions. To fully capture these suicide-related mental diatheses, we selected five relavent questionnaires containing altogether 34 subscales for clustering of children with STBs. five questionnaires are NIH cognitive battery ^48^, Child Behavior Checklist (CBCL) ^49,50^, Prodromal Psychosis Scale (PPS) ^51^, UPPS-P Impulsive Behavior Scale ^52^, and Behavioral Avoidance/Inhibition Scales (BIS/BAS) ^53^. Of all questionares/subscales : CBCL internalizing items reflects subjective distress, CBCL externalizing items, UPPS, BIS/BAS, and PPS scales reflects decision-making deficits, NIH cognitive battery characterizes cognitive deficits, and CBCL social problems reflects social distortions, see Supplemental Methods for details.

All 34 features underwent z-transformation before clustering. We used *eclust* function (visual enhancement of clustering analysis) in *factoextra* package in R v4.0 ^54^ to perform clustering analysis, which provide a framework automatically determining an optimal number of clusters (K) by gap statistic ^17^. K-means clustering with bootstrap adopting Euclidean distance was employed (K=2-10, maximum number of iterations=50, 100-time bootstrap, 80% resampling). Finally, the optimal K was used to produce the final clustering arrangement.

### Investiaging intergroup difference of STBs, psychiatric morbidity and potential risk factors across subtypes

For continuous variables (34 features) we used ANOVA (using *aov* function of R package *stat*; *TukeyHSD* function was used to do *post hoc* analyses). Covariates (sex, family income, parental education years, race, Hispanic, pubertal) were regressed out before ANOVA using *lm* function in *stat* package. For discrete variable, (STBs and psychiatric disorders), Chi-squared test (*chisq.test* function in *stat*) was performed and *chisq.posthoc.test* function was used *post hoc* analyses (*chisq.posthoc.test* package) ^55^.

### Examining multimodal neuroanatomical alterations in STB subtypes relative to HC

Multimodal brain imaging measures from gray matter, white matter and resting-state functional networks were compared between each subtype and HC to understand the underlying neural mechanisms of each STB subtype. For gray matter, we examined cortical thickness (CT) and cortical surface area (CSA) of 68 cortical regions (Desikan-Kiliany parcellation) ^56^ and volume of 14 subcortical regions (automatic subcortical segmentation) ^57^. For white matter, tract-averaged microstructural properties such as average fractional anisotropy (FA), mean diffusivity (MD), longitudinal diffusivity (LD), transverse diffusivity (TD) of 37 white matter tracts (AtlasTrack) ^58^ were analyzed, For functional networks, resting-state functional connectivity (FC) among 13 functional networks (from Gordon atlas) ^59^ was used. Details of preprocessing can be found in Supplemental Methods.

To investigate the cross-sectional and longitudinal (developmental) neuroimaging alterations of each subtype relative to controls, we estimated the inter-group main effect (compare each subtype with controls), development main effect (compare between age 10 and 12 for each group) and the group × development interaction effect using neuroimaging data. The linear mixed-effect models (R package *lmerTest*) were used to analyze the main and the interaction effects, which can control the site effects and were widely adopted in previous ABCD studies ^60,61^. Covariates were included as fixed effect [sex, race, body mass index (BMI), puberty, household income and parental education] or random effect variables (site and subject identity number) in LMMs. The model formula was as follows:

Brain Var = Intercept + β1 (Group) + β2 (Time Point) + β3 (Time Point *Group) + … βn (Covariates) + (1 | Subject) + (1 | Site) + *e*,

where Group is N-1 dummy variables (N is the optimal number of subtypes) where HC was treated as 0 and suicidal subtype as 1. For cortical and subcortical morphological analysis, whole brain volume was included as covariate; for FC, mean framewise displacement (FD) during scan was included; for DTI properties, average framewise motion was included. For each predictor, the LMM provides t-test results for the regression coefficient and a p value. FDR (q<0.05) was used for multiple comparisons.

## Data Availability

All data produced are available online at https://abcdstudy.org/

## Acknowledgements

LP acknowledges research support from the Canada First Research Excellence Fund, awarded to the Healthy Brains, Healthy Lives initiative at McGill University (through New Investigator Supplement to LP); Monique H. Bourgeois Chair in Developmental Disorders and Graham Boeckh Foundation (Douglas Research Centre, McGill University) and salary award from the Fonds de recherche du Quebec-Sante (FRQS). JZ was supported by Science and Technology Innovation 2030 - Brain Science and Brain-Inspired Intelligence Project (STI2030-Major Projects 2021ZD0200204), Shanghai Municipal Science and Technology Major Project (No.2018SHZDZX01), ZJ Lab, Shanghai Center for Brain Science and Brain-Inspired Technology, NSFC 61973086 and the State Key Laboratory of Neurobiology and Frontiers Center for Brain Science of Ministry of Education, Fudan University. XW was supported by State Key Laboratory of Medical Neurobiology and MOE Frontiers Center for Brain Science, and Institutes of Brain Science, Fudan University.

Data used in the preparation of this article were obtained from the Adolescent Brain Cognitive Development^SM^ (ABCD) study (https://abcdstudy.org), held in the NIMH Data Archive (NDA). This is a multisite, longitudinal study designed to recruit more than 10,000 children age 9-10 and follow them over 10 years into early adulthood. The ABCD Study® is supported by the National Institutes of Health and additional federal partners under award numbers U01DA0401048, U01DA050989, U01DA051016, U01DA041022, U01DA051018, U01DA051037, U01DA050987, U01DA041174, U01DA041106, U01DA041117, U01DA041028, U01DA041134, U01DA050988, U01DA051039, U01DA041156, U01DA041025, U01DA041120, U01DA051038, U01DA041148, U01DA041093, U01DA041089, U24DA041123, U24DA041147. A full list of supporters is available at https://abcdstudy.org/federal-partners.html. A listing of participating sites and a complete listing of the study investigators can be found at https://abcdstudy.org/consortium_members/. ABCD consortium investigators designed and implemented the study and/or provided data but did not necessarily participate in analysis or writing of this report. This manuscript reflects the views of the authors and may not reflect the opinions or views of the NIH or ABCD consortium investigators.

