## Supplemental Materials for "Identifying subtypes of youth suicidality based on psychopathology: alterations in genetic, neuroanatomical and environmental features"

**Supplemental Methods**

**Behavior and cognitive measures**

*Puberty.* A composite score for pubertal development was created by averaging across responses on three items related to general development and two sex-specific items and then averaging across parent- and child-report (range=1-4). These summary scores were averaged across parent and child report.

*Race.* The race was coded according to the demographic information of ABCD study and divided into 6 categories: "White", "Black", "AIAN/NHPI" (AIAN for American Indians and Alaskan Natives; NHPI for Hawaiian Natives and Pacific Islanders), "Mixed", "Asian" and "Others".

*Socio-Economic Status (SES).* The social economic status (SES) was measured by family income and years of education of parents. Income was the sum of the annual incomes of both parents, categorized as an ordinal variable across three bins (1: <$50,000; 2: $50,000-$100,000; 3: >$100,000). Years of education of parents were measured by the years of education of the parent with the highest education. Neighborhood Safety/Crime Survey (NSC). "< HS Diploma", "HS Diploma/GED", "Some College", "Bachelor", "Post Graduate Degree"

*NIH Cognitive Battery.* Cognitive abilities were measured by the NIH toolbox cognition battery (Casey et al., 2018; Weintraub et al., 2013), which included 7 tasks: picture vocabulary, flanker inhibitory control, picture sequence memory, card sort, pattern comparison processing speed, oral reading recognition and list sorting working memory. In addition, ABCD provided assessments of crystal intelligence, fluid intelligence, and total intelligence. Only 6 of 10 items are included in the 2-years-follow-up data (picture vocabulary, flanker inhibitory control, picture sequence memory, pattern comparison processing speed, oral reading recognition, crystal), $N_{follow-up2}$=6291.

*Kiddie Schedule for Affective Disorders and Schizophrenia Present and Lifetime Version (K-SADS).* Children and their parent/guardian completed modules of a self-administered, computerized Kiddie-Schedule for Affective Disorders (K-SADS COMP) (Kaufman et al., 1997; Townsend et al., 2020) to assess children’s lifetime (past, present, or partial remission) DSM-5 diagnoses (Barch et al., 2018). Composite variables were created to examine lifetime diagnoses of ADHD (parent-report only), depressive disorders (parent- or child-report of major depressive disorder, dysthymia, or an unspecified depressive disorder), anxiety disorders (parent- or child-report of separation anxiety disorder, social anxiety disorder, or generalized anxiety disorder), and externalizing disorders (parent-report of conduct or oppositional defiant disorder). In text, we examined the intergroup differences in 10 psychiatric disorders [attention deficit hyperactivity disorder (ADHD), oppositional defiant disorder (ODD), conduct disorder (CD), major depression disorder (MDD), general anxiety disorder (GAD), separation anxiety (SEP), Panic, social anxiety (SOC), post-traumatic stress disorder (PTSD), phobia, hallucination, delusion, bipolar disorder (BP), obsessive-compulsive disorder (OCD)]. All diagnoses were reported by K-SADS-P, coded as yes-no.

*Suicide diagnosis and suicide-related symptoms.* Suicide diagnosis refers to 23$^{rd}$ condition of DSM-5 ("Suicidal behavior disorder") from K-SADS-Parent and K-SADS-Youth. K-SADS-Parent included 17 suicide-related diagnoses and 23 suicide symptoms, while K-SADS-Youth included 20 suicide-related diagnoses and 22 suicide symptoms. Except for the two removed diagnoses in K-SADS-P and K-SADS-Y (“No Past suicidal ideation or behavior Past”, “No suicidal ideation or behavior Present”), all diagnoses and symptoms can be divided into four categories: self-injurious behaviors, suicidal ideation, suicidal attempts, and suicidal behaviors. Refer to SM Table 2 for specific classification.

*Child Behavior Checklist (CBCL) syndrome scales.* Parents/guardians completed the Child Behavior Checklist (CBCL) (T. M. Achenbach, 1999) to assess their child’s emotional and behavioral functioning. The 11 syndrome scales were calculated based on CBCL, including anxious/depressed, withdrawn/depressed, somatic complaints, social problems, thought problems, attention problems, rule-breaking behavior, aggressive behavior, internal problems, external problems, total problems (T. M. Achenbach, 1999; Ivanova et al., 2007) and 9 DSM-oriented scales (depression problems, anxiety problems, somatic problems, ADHD, oppositional defiant problems, conduct problems, sluggish cognitive tempo, obsessive-compulsive problems, stress problems) (T. Achenbach & Rescorla, 2007; T. M. Achenbach, Dumenci, & Rescorla, 2003). Age- and sex-normed T-scores were used for analyses. Parents of 11,870 children had completed CBCL at baseline assessment.

*Neighborhood Safety/Crime Survey (NSC).* The NSC survey was derived from the Neighborhood Safety/Crime measure in the PhenX Toolkit (Hamilton et al., 2011), evaluating the respondent's perception of safety and crime within their community. We utilized the average of the three NSC scores reported by parents as the measurement for NSC.

*Child Report of Behavior Inventory (CRPBI).* This study employed a condensed version of the Acceptance Scale from the Child Report of Behavior Inventory (CRPBI) (Barber & Olsen, 1997), selecting the top 5 items with the highest factor loadings from the original 10-item scale. The Acceptance Subscale assesses children's perceptions of caregiver warmth, acceptance, and responsiveness. It is divided into two parts, with the first section completed by the 'parent participant'—the adult responding to the parent surveys (variables labeled with 'mom,' even if not the mother). Subsequently, the survey is administered for a second primary caregiver (variables labeled with 'caregiver'), representing an adult with whom the child spends a significant amount of time (e.g., other parent, step-parent, grandparent). The total score serves as a comprehensive measure of acceptance.

*Family Environment Scale (FES).* The Conflict subscale of the Family Environment Scale (FES) (Roosa & Beals, 1990) comprises 9 items, assessing the frequency of open expression of conflict among family members. Both adolescents and parents provide responses to these questions.

*Parental Monitoring Survey (PMQ).* The Parental Monitoring Survey is employed to assess whether parents actively make efforts to monitor their children's activities both at home and when they are away, derived from two other measures (Karoly, Callahan, Schmiege, & Feldstein Ewing, 2016; Stattin & Kerr, 2000).

*Sleep Disturbance Scale (SDS) for Children.* The questionnaire assesses sleep disturbance in children during the sleep process, including factors such as insomnia, nocturnal awakenings, and etc.(Bruni et al., 1996).

*Sports and Activities Involvement Questionnaire.* Involvement in sports, music, hobbies, art, theater, reading, and TBI risk. Parents were asked for how many years their child had participated in a given activity. The maximum allowed for this question was 10 years.

*School Risk and Protective Factors (SRPF).* The School Risk and Protective Factors (SRPF) survey is derived from the PhenX School Risk and Protective Factors protocol, adapted from the " The Communities That Care (CTC) Youth Survey " (Arthur et al., 2007). The SRPF survey assesses youths' perceptions of the school atmosphere and involvement, also requesting them to report on academic performance. The responses are utilized to derive scores for three subscales: School Environment, School Engagement, and School Disengagement.

*Youth Screen Time Survey.* This measurement includes customized questions regarding the total time adolescents spend on visual media on both weekdays and weekends. The assessed media activities encompass: (1) watching TV shows or movies; (2) viewing videos (such as YouTube); (3) playing electronic games on computers, gaming consoles, smartphones, or other devices; (4) texting on phones, tablets, or computers; (5) accessing social networking sites like Facebook, Twitter, Instagram; (6) video chatting. There are seven response options: none, <30 minutes, 30 minutes, 1 hour, 2 hours, 3 hours, 4 hours or more. In this study, the total time spent engaging in various screen activities throughout the week is aggregated as the overall score for children's screen time across different activities.

**Neuroimaging measures and quality control**

*Structural Magnetic Resonance Imaging (sMRI).* All brain imaging data was acquired on GE, Siemens, or Phillips scanners. High-resolution T1- and T2-weighted structural Magnetic Resonance Images (1mm isotropic, prospective motion correction) of all children were collected, and processed through ABCD pipelines (mainly including registration, intensity normalization and bias field correction) (Casey et al., 2018). Cortical volume was obtained from T1-weighted structural images, segmented and reconstructed by Freesurfer 5.3 (Fischl, 2012). Subcortical structures are labelled using an automated, atlas-based, volumetric segmentation procedure (Fischl et al., 2002). Labels for cortical gray matter were assigned based on Desikan parcellation, which divides brain cortex into 68 regions of interest (ROIs) (Desikan et al., 2006).

*Diffusion Tensor Imaging (DTI).* High angular resolution diffusion imaging (1.7mm isotropic) was collected using multiband acquisition (b-values=4, factors=4, directions=96) (Casey et al., 2018), then a standardized preprocessing included eddy current, head motion, and distortion correction was performed (Hagler Jr et al., 2019). Major white matter tracts were automatically segmented using AtlasTrack (Hagler Jr et al., 2009). ABCD study provided common DTI parameters such as average factional anisotropy (FA), mean diffusivity (MD), longitudinal diffusivity (LD), transverse diffusivity (TD) as well as Restriction Spectrum Imaging (RSI) model parameters (White, Leergaard, D'Arceuil, Bjaalie, & Dale, 2013) of 37 main tracts.

*Resting-State Functional Connectivity (RSFC).* High-resolution resting-state functional MR imaging of children were acquired by multi-band scanning (2.4 mm isotropic, TR=800ms, 6 factors). Standardized fMRI preprocessing included registration, distortion correction, and normalization. Post-processing included regression of 24 temporally filtered motion parameters, frame-wise displacement (FD)>0.3mm outliers, as well as white matter, cerebral spinal fluid, and whole brain signal (Hagler Jr et al., 2019). Within- and between-network connectivity was extracted by averaging all connections between ROIs assigned to given networks of the Gordon atlas (Gordon et al., 2016).

*Quality Control of Imaging Data.* Par quality control (QC) information provided by QC file (*abcd_imgincl01*) from ABCD 3.0 release, data quality control was performed for all brain imaging data according to the quality control parameters recommended by ABCD study (Hagler Jr et al., 2019).

**ABCD follow-up data**

The ABCD V3.0 dataset had released part of data of the following two years of follow-up. The 1-year follow-up included 11,235 children of 11,878, and the 2-year follow-up included 6,571 children.

Follow-up differed for each type of data. Among the data used in this study, brain imaging data and K-SADS-P included two time points, baseline and 2-year follow-up, only, while K-SADS-Y included three time points, baseline, 1-year, and 2-year follow-up.

**Validation analysis**

Based on gap statistics, the same optimal K values (K=5) were obtained on both sets of data in the split-half validation (SM Fig 2). The clustering results of spilt-half validation were similar to those of all 1,624 subjects, including one significant depressive subtype, one externalized subtype, and one high-functioning subtype. Subtypes 3 and 4 differ slightly in both parts of the data, but still show stable traits overall (subtype 3 has lower intelligence and subtype 4 has higher psychotic prodrome and impulsivity). See SM Fig 3.

We performed separate clustering based on parent- and youth-reported suicides among children. Clustering using parent-reported cases identified five subtypes, including that the subtypes of 1, 2, 3, 5 found in the main analysis, and a moderate depression subtype (more depressive than subtype 2, less than subtype 1) or subtype 6, while clustering using youth-reported cases identified all five subtypes of preexisting and a similar subtype 6 with moderate depression (see SM Fig 4 and SM Table 15). This clustering result shows that our clustering result has high stability.

After adjusting for the effects of crystal intelligence, the volume of subtype 1-4 still showed decreasing trend [Subtype 1 ($t_{4239.5}$=-2.53, p=1.13e-2), Subtype 2 ($t_{4219}$=-5.56, p=2.83e-8), Subtype 3 ($t_{4246.2}$=-3.298, p=1e-3), Subtype 4 ($t_{4207.5}$=-2.82, p=4.89e-3)], while the increase of total brain volume of Subtype 5 disappeared ($t_{4237.5}$=1.87, p=6.2e-2>0.05). In terms of local brain indices, the increasing cortical thickness of Subtype 5 disappeared. Subtype 2 no longer showed significant functional connectivity abnormalities. Subtype 3 still showed abnormalities in the white matter fiber tracts, but the area was significantly reduced. See SM Fig 5.

### Supplemental Figure

### SM Figure 1. Comorbidity of suicidal children

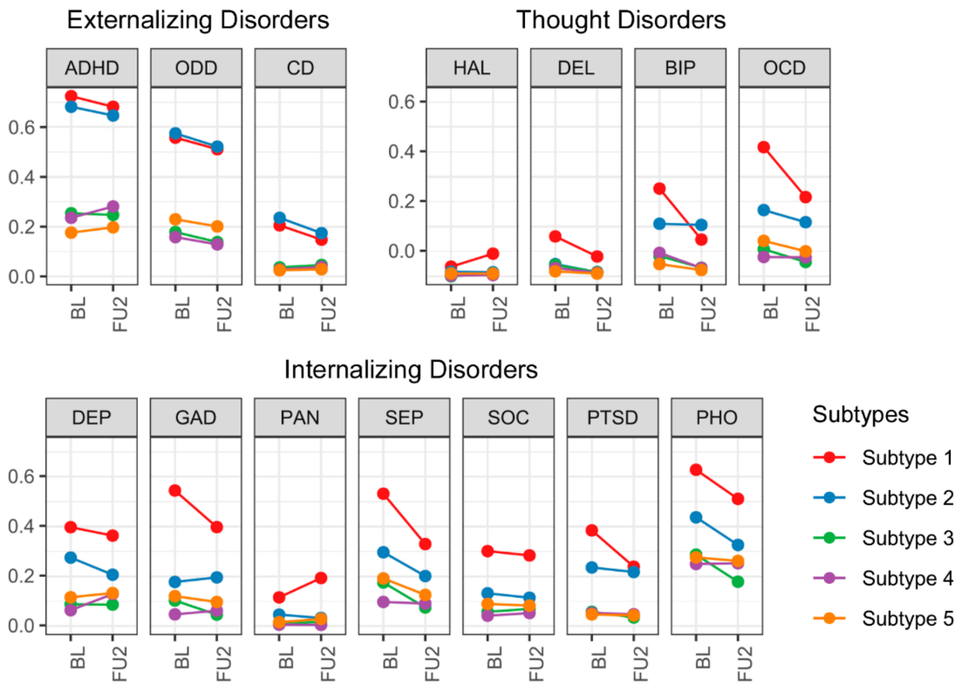

260 **SM Figure 2. Gap statistics of spilt-half validation**

261 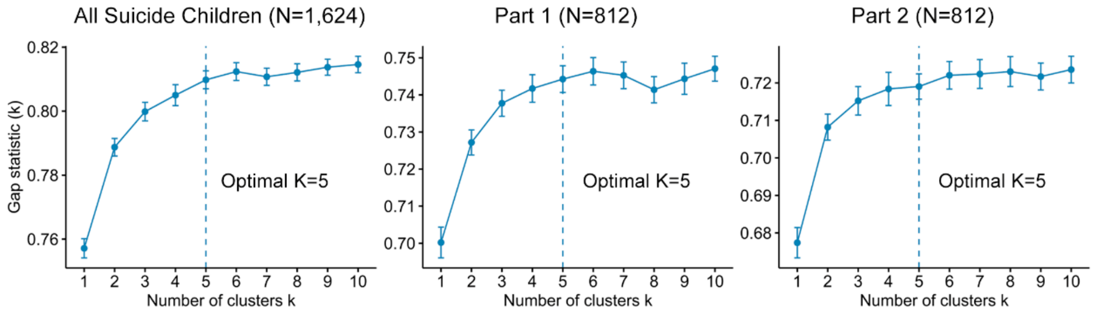


262

263 **SM Figure 3. Clustering result of spilt half validation**

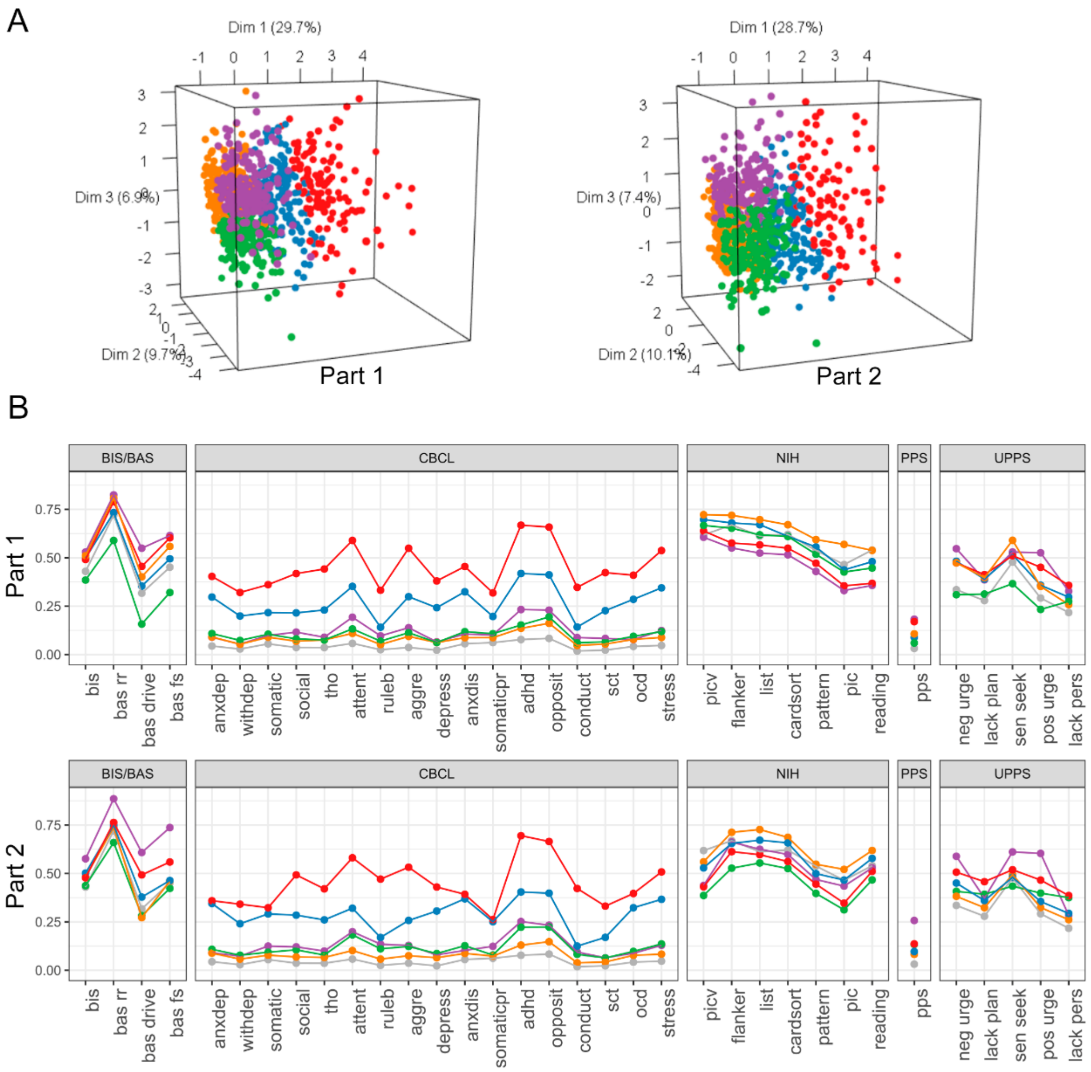


264

265

266

**SM Figure 4. Clustering results from cases separate defined by K-SADS-P & K-SADS-Y**

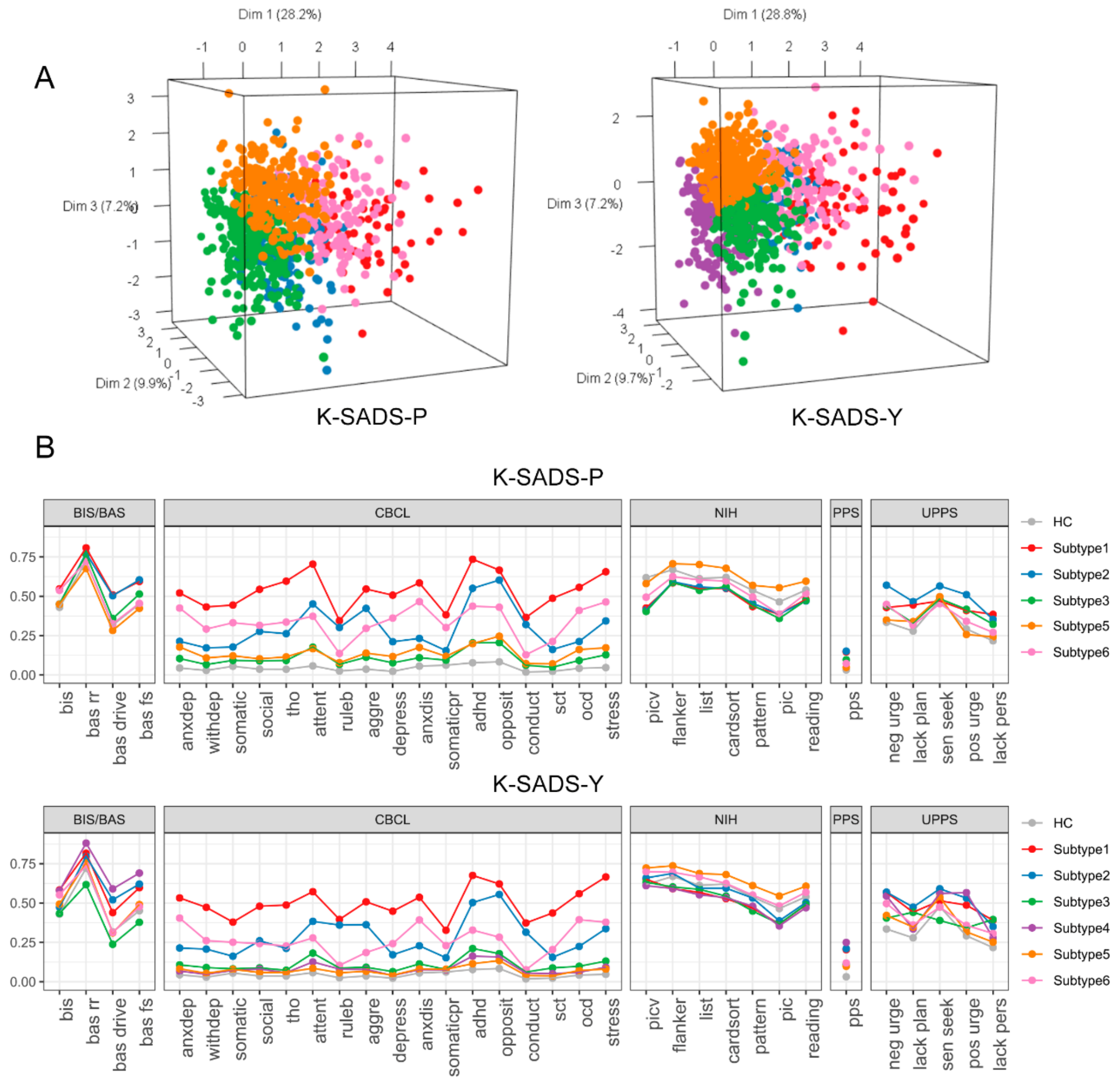

**SM Figure 5. Brain Intergroup difference (regressing crystal intelligence out)**

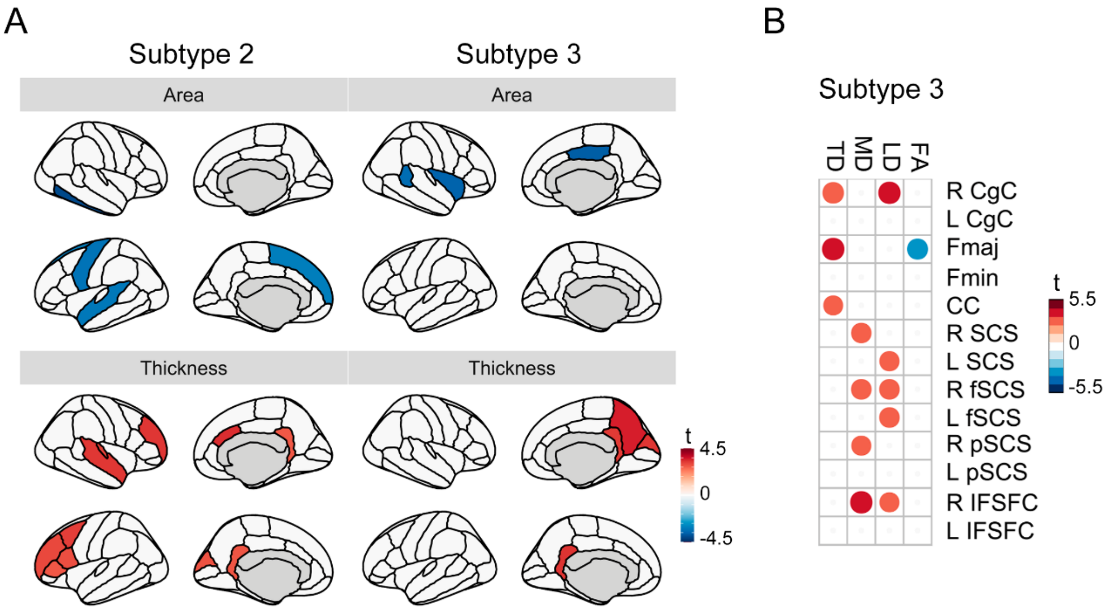

**Supplemental Table**

**SM Table 1 Questionnaire List.**

**SM Table 2 The centroids of 5 child suicide subtypes obtained through cluster analysis.** The displayed values represent the mean scores of z-standardized questionnaire responses across different subtypes.

**SM Table 3 Suicide thought and behaviors and psychiatric morbidity profile of 5 subtypes.** The values shown in the table represent the number of individuals within each subtype who have specific mental disorders or exhibit particular suicidal thoughts or behaviors. The numbers in parentheses indicate the proportion of these individuals relative to the total population of that subtype. The last column displays the p-values from the chi-square test. Morbidity: ADHD, Attention Deficit Hyperactivity Disorder; ODD, Oppositional Defiant Disorder; CD, Conduct Disorder; MDD, Major Depression Disorder; GAD, General Anxiety Disorder; SEP, Separation Anxiety; SAD, Social Anxiety; PTSD, Post-Traumatic Stress Disorder; BIP, Bipolar Disorder; OCD, Obsessive-Compulsive Disorder.

**SM Table 4 Suicide thought and behaviors and psychiatric morbidity profile (posthoc analysis of Chi-square test).** The residual column represents the difference between the observed and expected values for each cell in the contingency table. The p-value indicates significance, and when $p < 0.05$ (FDR corrected) for a subtype, it signifies that the proportion of a specific symptom in that subtype significantly differs from the other subtypes.

**SM Table 5 Intergroup difference of diatheses and risk factors (ANOVA).** The table displays the F-value, p-value, and degrees of freedom (DF) for the ANOVA analysis, respectively.

**SM Table 6 Intergroup difference of diatheses and risk factors (ANOVA post-hoc analysis).** The table presents the z-value differences (diff) and p-values for pairwise comparisons between each group (5 subtypes and HC).

**SM Table 7 sMRI Results.** The table displays the t-values, degrees of freedom (DF), and p-values corresponding to the brain changes in each subtype relative to HC.

**SM Table 8 rs-fMRI Results.** The table displays the t-values, degrees of freedom (DF), and p-values corresponding to the brain changes in each subtype relative to HC.

**SM Table 9 DTI tract Results.** The table displays the t-values, degrees of freedom (DF), and p-values corresponding to the brain changes in each subtype relative to HC.

**SM Table 10 PRS (ANOVA).** The table displays the F-value, p-value, and degrees of freedom (DF) for the ANOVA analysis.

**SM Table 11 PRS (ANOVA post-hoc analysis).** The table presents the z-value differences (diff) and p-values for pairwise comparisons between each group (5 subtypes and HC).

**SM Table 12 Longitudinal STBs.** The table presents the proportions of self-harm, suicidal ideation, suicide attempts, and suicide behavior exhibited by each subtype at baseline (BL, 10 years old), 1-year longitudinal follow-up (FU1, 11 years old), and 2-year longitudinal follow-up (FU2, 12 years old).

**SM Table 13 Results of repeated sample ANOVA of Longitudinal STBs.** The table presents the F-values, degrees of freedom (DF), and p-values for the subtype main effect, time main effect, and subtype-by-time interaction effect of suicide scores (STB score) across various subtypes from baseline to a 2-year follow-up. Furthermore, the table elaborates on the interaction effect, depicting the magnitude of age-related changes in STB scores for each subtype relative to all other groups.

**SM Table 14 Paired comparison results of repeated sample ANOVA of Longitudinal STBs.** The table illustrates the results of pairwise comparisons for the temporal trends in STB scores across various subtypes.

**SM Table 15 Validation Results of Spilt-Half Analysis and Clustering in K-SADS-P/K-SADS-Y only.** The table presents the results of split-half validation for cluster reliability and individual validation using K-SADS-P/K-SADS-Y. Reliability is measured based on the correlation coefficients between the clustering centroids obtained from the validation sample and those from the entire sample. The r-value represents the Pearson correlation coefficient, and p represents the corresponding p-value of the correlation coefficient.

**SM Table 16 Gap Statistics.** The table displays the Gap Statistics used to determine the optimal number of clusters in the clustering analysis. For each cluster count (k), the gap calculation compares the total within-cluster deviation sum log(W(k)) of the original data at different k with the expected within-cluster deviation sum E*[log(W(k))] under the assumption of a uniform distribution.
